# Correlates of functional physical capacity in physically active older adults: a conceptual-framework-based cross-sectional analysis of social determinants of health and clinical parameters

**DOI:** 10.1101/2022.02.02.22270171

**Authors:** Lucinéia Orsolin Pfeifer, Lucas Helal, Nórton Luís Oliveira, Daniel Umpierre

## Abstract

This study aimed to explore social determinants of health and health/clinical determinants on two outcomes of functional physical capacity. Therefore, a population-based sample of 327 older adults (69±7 years; 83.5% women) underwent demographical and clinical questionnaires, risk factors assessments, six-minute walk testing (walking capacity), and handgrip strength testing. Based on multivariable linear regression models, age (−4.05m; - 5.3 to −2.8), being men (71.40m; 50.5 to 92.3), body mass index (−3.88m; −5.6 to −2.1), and quality of life (18.48m; 6.3 to 30.6) remained as predictive variables for walking capacity (R^2^=30.8%). In the final model for handgrip strength, age (−0.6% kgf; 0.89 to 0.2) and male sex (65.2% kgf; 55.3 to 75.8) remained as predictive variables. Despite exploratory analyses including contextual factors as potential predictors of walking capacity and handgrip strength, only outcomes at the individual levels were associated, either positively or negatively, with the variations presented by this studied sample of older adults.

## INTRODUCTION

Functional decline in older adults may be affected by clinical status, physical fitness, and social determinants of health (Beaton et al., 2015). In this regard, sarcopenia, which is a progressive decline in skeletal muscle mass (Evans, 1995), has been consistently associated with a decline in functional capacity and disability in the elderly. Similarly, dynapenia, which relates to the loss in muscle strength independently of changes in muscle mass is also an independent predictor of overall mortality in this population (Clark & Manini, 2012). In terms of functional capacity outcomes, handgrip and lower limb muscle strength, and ambulatory gait speed are indexes of physical fitness and may predict adverse events in older adults, such as declines in cognition, mobility, functional status (Rijk et al., 2016), disability, (Donoghue et al., 2014; Shimada et al., 2015), hospitalization (Cesari et al., 2009) and all-cause mortality (De Buyser et al., 2013; Rantanen et al., 2003; Rijk et al., 2016; Studenski et al., 2011). Importantly, regular exercise may counteract these deleterious effects (Izquierdo et al., 2021), both by structured exercise training and physical activity advice, thus optimizing the maintenance of functional capacity during aging.

Similar to the many health-related events, the functional decline in older adults is potentially influenced by a combination of social determinants of health and health/clinical-related determinants. Age, sex, and body mass have a solid association with ambulatory functional outcomes (i.e., impaired walking test and handgrip strength) (ATS Committee on Proficiency Standards for Clinical Pulmonary Function Laboratories, 2002; Casanova et al., 2011; Corrêa et al., 2020; V. Z. Dourado, 2011; Gale et al., 2007; Lu et al., 2020; Salbach et al., 2015; Tveter et al., 2014) and are frequently used in trials as surrogate outcomes (V. Z. Dourado, 2011). Although some potential predictors more income, use of medications, diabetes, quality of life, and depression are, likewise, associated (Enright et al., 2003; Kuziemski et al., 2019; Liang et al., 2020; Ling et al., 2010; Love et al., 2020; Serra et al., 2015), although not explored as far as the other potential predictors aforementioned. The variability in results of functional tests can be explained through a myriad of factors (standards of the tests, color, ethnicity, and geographic locations, even in the same country), which make the inter and intra-test comparability inappropriate to other populations.

We perform out the first exploratory study from a sample initially evaluated in a cross-sectional descriptive study carried out previously (F. M. Dourado et al., 2021). We aimed to estimate, in a representative sample of older adults engaged in a municipal program of physical activity, the extent of the contribution of social and clinical health determinants in functional physical capacity, here defined as the ambulatory walking capacity and the handgrip force. As this is an exploratory study, we do not present a directional hypothesis.

## METHODS

### Study setting

This is the first exploratory study within the descriptive cross-sectional study entitled: *“Health profile of elderly users of a town public program of physical activity: a cross-sectional study”*. In this study, we used questionnaires, clinical measurements, and functional tests to assess the health profile and social determinants of health of older adults spontaneously engaged in a public lifestyle program composed of several lifestyle items, including a physical activity program, in the city of Porto Alegre, Brazil. In the city public program, modalities such as gymnastics, resistance training, rhythms, and/or recreational sports were offered to participants. The majority of them took place twice a week lasting one hour of duration (F. M. Dourado et al., 2021). The study protocol is publicly available at https://osf.io/q4r69/ and was approved by the Institutional Review Board of the Hospital de Clínicas de Porto Alegre (GPPG 2018-0100; CAAE 84093317600005327) and of the Municipal Secretariat of Health (SMS/POA). The procedures followed national guidelines for bioethics in research. All participants read and signed the informed consent before starting their enrollment in the study. The reporting of this manuscript is based on the STROBE Statement recommendations (Strengthening the reporting of observational studies in epidemiology) in its extension for cross-sectional studies (von Elm et al., 2007). Study data are also available in a public repository for independent researchers (https://zenodo.org/record/4341443#.X9u61thKhPY; doi: 10.5281/zenodo.4341443).

### Participants and eligibility criteria

The inclusion and exclusion criteria defined in the previous descriptive study were: being 60 years old or older and regularly attending the physical activity programs of the municipal community centers. Exclusion criteria were defined as the impossibility of moving to the assessment sites and/or any musculoskeletal injury which put the patient at risk in any setting. All participants included in the previous study were considered for inclusion in this study provided they had complete assessment data.

### Sampling and sample size calculation

The sample size calculation was carried out for the previous descriptive study, using the prevalence of hypertension as the outcome of interest and considering the size of each of the centers in the city, totaling 443 participants. Detailed information is described in the previous study (F. M. Dourado et al., 2021).

### Data collection

The recruitment and data collection were carried forward between April 2018 and February 2019. Data were collected at two moments. In visit 1, the research team went to 11 different sites attended by the participants. The data collection included self-administered questionnaires and the 6-minute walking test (6MWT). In visit 2, the participants went to the Hospital de Clínicas de Porto Alegre, Clinical Research Center, undergoing clinical measurements (e.g., blood pressure readings and anthropometric evaluation) and handgrip testing. The procedures for data collection are more detailed elsewhere (F. M. Dourado et al., 2021).

The researchers received training in the standardization of procedures; some of the measurements were routinely performed by the research team for other studies (Umpierre et al., 2019), based on a manual of standard operating procedures. This was done to minimize measurement bias since different researchers were involved in the measurements and procedures in different centers visited.

### Study Variables

### Social determinants of health and clinical/health-related variables

A 62-item questionnaire was applied (visit 1), allowing the collection of demographics, clinical history, social determinants of health, risk factors, diet, physical activity behaviors, morbidity, and perception of health. Sex was categorized into men and women; the number of medications in four categories (none, up to 3 medications, 4 to 6 and 7 or more); diabetes as yes, no, or gestational; family income in four minimum wages (MW) categories (up to 2 MW, 2 to 4 MW, 4 to 10 MW, 10 to 20 MW).

The older adult’s height (cm) and weight (kg) were transformed into body mass index (BMI) values by taking the weight in kilograms and dividing it by the square of height in meters (kg/m^2^).

Quality of life was assessed using the Short Form 6 Dimension (SF-6D) self-administered questionnaire, which consists of six questions, within six domains, namely: functional capacity, physical and emotional aspects, social aspects, pain, mental health, and vitality. The questionnaire generates a score between 0.29 - 1.00, whether 1.00 means a full quality of life state.

Depressive symptoms were assessed by the Geriatric Depression Scale (GDS-15), which consists of 15 questions. In GDS-15, each positive answer associated with depression represents a point, generating scores from 0 to 15, where scores of 6 or higher were used as suggestive of depression (Krishnamoorthy et al., 2020). Using this cutoff point (score 6), the variable was categorized into presence or absence of depression.

#### Functional Capacity

##### 6MWT

The 6MWT was used to indirectly assess the cardiorespiratory fitness (estimations) and walking capacity. We considered the distance covered during a 6-minute period, in a 30-meter delimited pathway. The participants were encouraged in a standardized manner, according to the established guidelines (ATS Committee on Proficiency Standards for Clinical Pulmonary Function Laboratories, 2002).

##### Handgrip Test

The handgrip strength test was used to measure the strength generated by the muscles of the hand and forearm, and assess the physical condition of the upper limb, an important proxy of health status. Three measurements were taken using a dynamometer (Jamar, model 2A, Asimow Engineering Co., Santa Monica, USA), at a standing position of participants, and the elbow at 90 degrees of flexion. We took a series of measurements for both hands and intervals of one minute were given between each attempt. For the purpose of this study, the highest value reached in kgf was considered the dominant hand.

### Statistical methods

Continuous variables are described as mean and standard deviation whereas categorical variables are reported as prevalences and absolute frequencies. Most variables were maintained in the way they were collected, however, some variables were categorized to facilitate data analysis and interpretation (i.e., number of medications, and family income). We evaluated distributions visually and statistically - the first through a visual inspection of histograms and Kolmogorov-Smirnov tests which assume normality of the data as H0.

For inferential analysis, we used a multivariable linear regression model. Components of functional capacity were defined as dependent variables for this study in two separate models. To maintain similarities between analyses and preserve the natural units of the coefficients, we transformed the handgrip observations into a natural logarithm (log), which generated a Gaussian distribution and allowed us to carry on a linear regression model. Then, we regressed both log of handgrip strength and 6WMT by linear multivariable regression analysis using a Gaussian distribution for the outcome and an identity function as linkage, in a two-step manner (model 1 and model 2 - the last one adjusted to more variables based on our conceptual framework model - explained below).

In regression analyses, we diagnosed the basic assumptions of multivariable linear regressions (i.e., normality of the distribution of the outcome and its residual, multicollinearity, and constant variance of residuals), the adequacy of the chosen model predictors the model’s effectiveness. We plotted adjusted outcomes against Pearson residuals and independent variables against residuals, as well as a tested correlation between them and the assumption of homoscedasticity. When exploring heterogeneity, we found evidence of heterogeneity of variances, mostly due to observations in sex, about the dependent variable handgrip. Based on this, we carried out a linear model for 6WMT and a linear model using Generalized Least Squares (GLS) for the log handgrip strength.

The variables included in the multivariable linear regression model were defined *a priori* based on a causal conceptual framework. Distal and proximal variables were inserted according to the knowledge base (e.g., obtained through published literature) for potential causality. We chose the conceptual framework method because stepwise models may have limitations if no causal relationship is considered. These models may have limited power to select important predictors and correlates in small datasets and may result in a bias regarding the estimated regression coefficients (Steyerberg et al., 1999). Because thresholds of statistical significance are limited as a selection method to determine which variables should be added to a regression model(Bagherzadeh-Khiabani et al., 2016), our variable selection model for inclusion in the regression model is based on identified direct evidence of a possible association and physiological plausibility relationship of the influence of covariates. Considering that model 1 of the regression was composed of the proximal variables, which were loaded into model 2 (distal variables) only when they reached a correlation with the dependent variable (*P*<0.05).

No data imputation was done. In this analysis model, only the complete data of all variables present in the regression model remained, that is, any participants that presented missing data were excluded. To claim statistical significance, we assumed a threshold of 0.05. Analyses were performed in the software SPSS (Statistical Package for the Social Sciences) version 18.0 and R (R Core Team 2020, version 4.0.3). R: A language and environment for statistical computing. R Foundation for Statistical Computing, Vienna, Austria. R Packages used were tidyverse, nlme, emmeans.

## RESULTS

In total, 327 participants were included in the current study. Participants had a mean age of 69±7 years (min to max 60 up to 91 years old), and 83.5% of them were female (**Table 1**). The mean BMI was 27±3 kg/m^2^ and 47.4% of the participants were classified as “overweight”. The prevalence of self-reported diabetes (any type) was 12.5%. Potential depression, as classified by the depression scores of the GDS-15 instrument, had a prevalence of 18%; and, importantly, QoL levels had a mean score of 0.87±0.06. Concerning family income, 73.2% of participants received up to 4 MW. 70 participants (21.4%) reported not using any type of continuous-use medication, although the vast majority took at least three medications continuously as prescribed. Commenting on our primary outcomes (dependent variables), the mean walk capacity achieved was 498.4±78 and the mean handgrip strength by the dominant hand was 27.1 kgf±8.2 kgf (min to max from 302m up to 690m and 8 kgf to 60 kgf, respectively for both outcomes). **Table 2** shows the descriptive analysis of dependent variables and predictors.

**Table 1.**
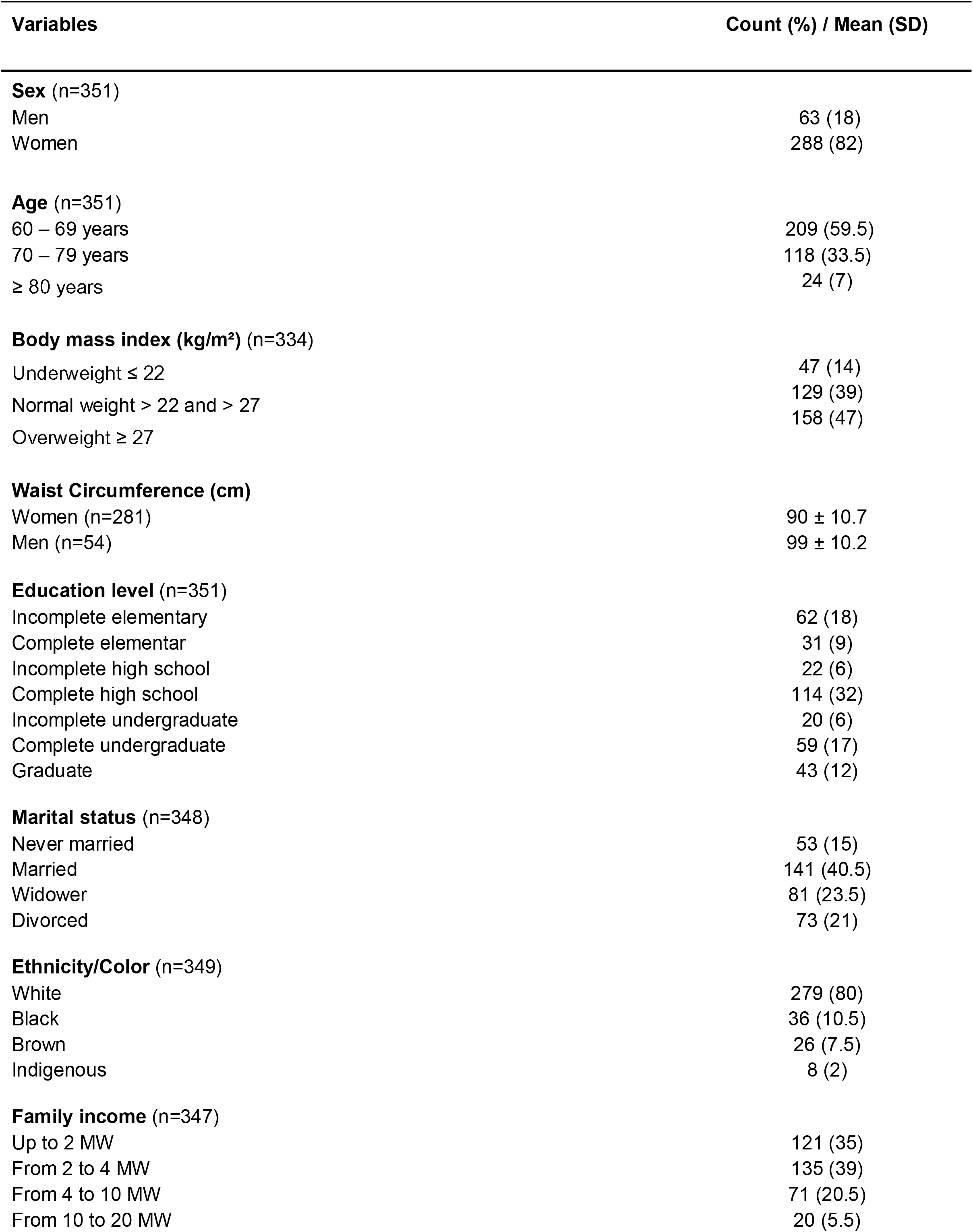
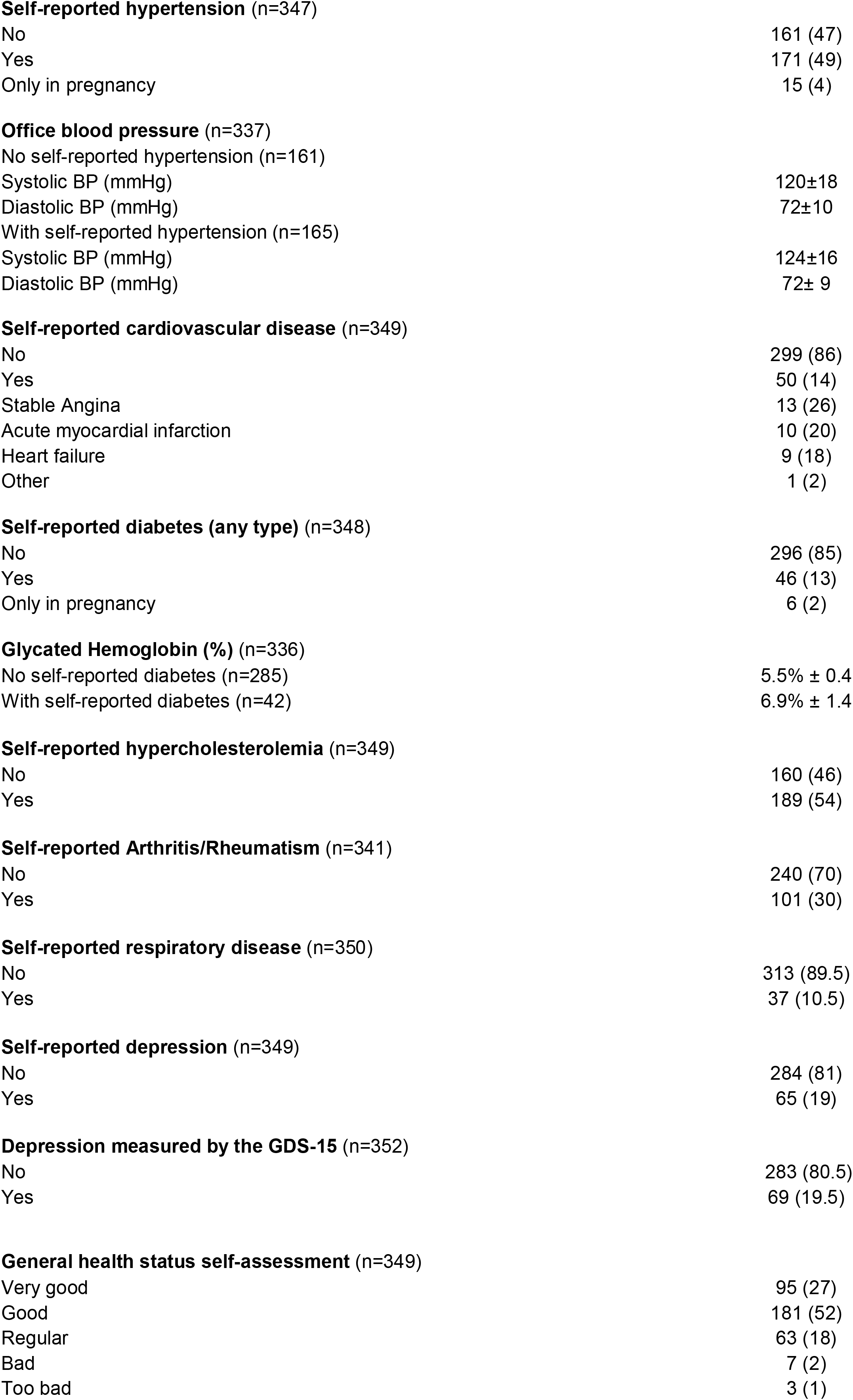
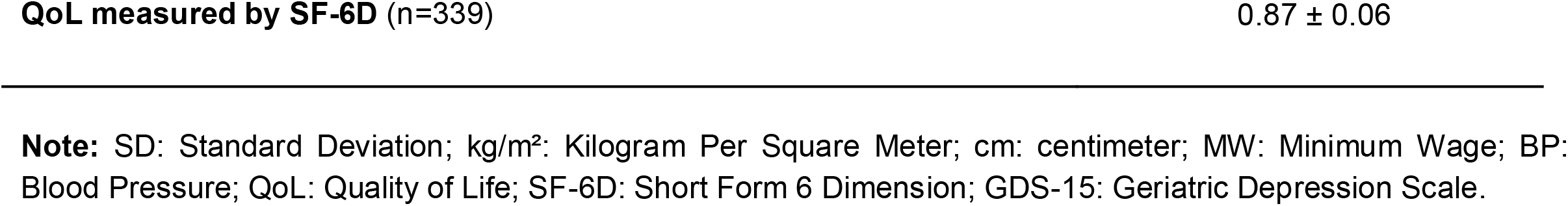
Baseline characteristics of all participants.

**Table 2.** Dependent variable with the respective categorical variables used as predictors in each analysis.

| Outcomes | Prevalence (%) | Mean 6WMT | CI 95% 6WMT |
| --- | --- | --- | --- |
| **6WMT(m) (n=312)** |  | 500.0 | (491.1 - 508.4) |
| **Sex** |  |  |  |
| Female (n=262) | 84.0 | 488.4 | (479.7 - 497.1) |
| Male (n=50) | 16.0 | 559.1 | (536.1 - 582.1) |
| **Number of medications** |  |  |  |
| None (n=69) | 22.0 | 515.6 | (497.0 - 534.1) |
| Up to 3 (n=149) | 48.0 | 501.5 | (489.7 - 513.4) |
| From 4 to 6 (n=79) | 25.2 | 483.1 | (466.5 - 499.8) |
| 7+ (n=15) | 4.8 | 463.9 | (430.3 - 497.6) |
| **Diabetes diagnosis** |  |  |  |
| No (n=266) | 85.3 | 503.4 | (494.0 - 512.9) |
| Yes (n=40) | 12.8 | 476.0 | (452.4 - 499.6) |
| Gestational (n=6) | 1.9 | 495.3 | (460.3 - 530.4) |
| **Family income** |  |  |  |
| Up to 2 MW (n=109) | 35.0 | 479.6 | (466.6 - 492.6) |
| 2 to 4 MW (n=118) | 37.8 | 499.1 | (484.2 - 513.9) |
| 4 to 10 MW (n=65) | 20.8 | 525.4 | (508.3 - 542.5) |
| 10 to 20 MW (n=20) | 6.4 | 530.7 | (489.5 - 571.8) |
| **Depression - GDS-15** |  |  |  |
| No (n=257) | 82.4 | 504.9 | (495.6 - 514.2) |
| Yes (n=55) | 17.6 | 475.8 | (453.7 - 497.9) |

|  |  | Mean Handgrip | 95%CI Handgrip |
| --- | --- | --- | --- |
| **Handgrip (kgf) (n=325)** |  | 27.0 | (26.2 - 28) |
| **Sex** |  |  |  |
| Women (n=271) | 83.4 | 24.5 | (24.0 - 25.1) |
| Men (n=54) | 16.6 | 40.0 | (37.8 - 42.3) |
| **Diabetes diagnosis** |  |  |  |
| No (n=279) | 85.9 | 27.2 | (26.2 - 28.2) |
| Yes (n=40) | 12.2 | 26.5 | (24.0 - 29.0) |
| Gestational (n=6) | 1.9 | 25.2 | (22.4 - 28.0) |
| **Number of medications** |  |  |  |
| None (n=69) | 21.3 | 27.8 | (25.8- 29.8) |
| Up to 3 (n=156) | 48.0 | 26.6 | (25.4 - 27.9) |
| From 4 to 6 (n=83) | 25.5 | 27.8 | (25.9 - 29.7) |
| 7+ (n=17) | 5.2 | 24.9 | (21.3 - 28.5) |

**Note:** 6MWT: 6-minute walk test; kgf: kilograms/force SE: Standard Error; CI: Confidence Interval; MW: Minimum Wage; GDS-15: Geriatric Depression Scale.

We regressed the 6MWT in a multivariable linear regression of 312 participants (**Table 3**). Model 1, including predictors as (age, sex, BMI, number of medications, and quality of life (SF-6D)) explained 30% (adjusted R^2^) of the variability in walking distance. For each unit of age (*P*<0.001), BMI (*P<*0.001), sex (*P*<0.001), and QoL (*P*=0.001), which remained independent correlates, modified the covered distance in 6 minutes about a decrease of - 4.04m (−5.2m to −2.7m), −4.30m (−6.0m to −2.6m) and 72.84m (52.9m to 92.8m), respectively; and, the QoL increases, the 6MWT increases, in average, 20.58m (8.9m to 32.2m). In a second model, we maintained variables with statistical significance and added into the model 1 potential predictors/correlates as diagnosis of diabetes, family income, and potential depression. The addition of such variables roughly modified the variance (adjusted R^2^) = 30.8% vs 30%). All previous independent variables remained correlated with 6WMT and the new inserted variables did not reach correlation to 6MWT.

**Table 3.** Multivariable linear regression between 6MWT test, social determinants of health and clinical parameters.

| **Model 1:** adjusted analysis for all predictors below |  |  |  |
| --- | --- | --- | --- |
| **Predictors (N=312)** | **$\beta$** | **CI 95%** | **P-value** |
| Age | -4.04 | -5.2 to -2.7 | <0.001 |
| Sex (male) | 72.84 | 52.9 to 92.8 | <0.001 |
| BMI | -4.30 | -6.0 to -2.6 | <0.001 |
| Number of medications (up to 3) | -0.56 | -19.2 to 18.1 | 0.95 |
| Number of medications (from 4 to 6) | -3.34 | -24.7 to 18.0 | 0.76 |
| Number of medications (7+) | -17.51 | -55.0 to 20.0 | 0.36 |
| QoL (SF-6D) | 20.58 | 8.9 to 32.2 | 0.001 |
| $R^2 = 0.32$; Adjusted $R^2 = 0.30$ | | | |
| **Model 2:** adjusted analysis for significant variables in model 1 plus diabetes diagnosis, family income and depression |  |  |  |
| Age | -4.05 | -5.3 to -2.8 | <0.001 |
| Male sex | 71.40 | 50.5 to 92.3 | <0.001 |
| BMI | -3.88 | -5.6 to -2.1 | <0.001 |
| QoL (SF-6D) | 18.48 | 6.3 to 30.6 | 0.003 |
| Diabetes diagnosis (yes) | -14.05 | -70.6 to 42.5 | 0.63 |
| Diabetes diagnosis (no) | 4.40 | -48.6 to 57.4 | 0.87 |
| Family income (up to 2 MW) | -13.53 | -45.8 to 18.7 | 0.41 |
| Family income (2 to 4 MW) | -17.19 | -48.6 to 14.2 | 0.28 |
| Family income (4 to 10 MW) | -4.22 | -37.5 to 29.0 | 0.80 |
| Depression - GDS-15 (no) | 16.56 | -4.0 to 37.1 | 0.11 |
| $R^2 = 0.33$; Adjusted $R^2 = 0.308$ | | | |

**Note:** CI: Confidence Interval; BMI: Body Mass Index; QoL: Quality of Life; SF-6D: Short Form 6 Dimension; MW: Minimum Wage; GDS-15: Geriatric Depression Scale. The reference category for sex: female; number of medications: no medications; diabetes diagnosis: gestational diabetes; family income: 10 to 20 minimum wages and for depression: yes.

To the handgrip strength, the results of the analysis of multivariable regression on generalized linear models with 325 participants are presented in **Table 4**. In the first model, age, sex, BMI, and the number of medications were included according to our conceptual framework. For each unit of age (*P*=0.009), sex (male) (*P*<0.001) and use of seven or more medications (*P*=0.02), which remained independent correlates, modified handgrip performance in an average decrease of −0.5 kgf% (0.89 kgf to 0.09 kgf), and −13.1 kgf% (22.74 kgf to 2.17 kgf), for age and seven or more medications, respectively; and a 65.9 kgf (55.89 kgf to 76.47 kgf) increase for males, in average. In model 2, we maintained variables with statistical significance from model 1 and added diagnosis of diabetes. Only the independent variables age (*P*=0.005) and sex (*P*<0.001) remained correlated with the handgrip test, the number of medications (seven or more) lost correlation (*P*=0.07) and the newly inserted variable did not reach a correlation with the test. However, age and sex (men) were practically not impacted either after adjustment for the additional variables, neither their precision estimators −0.5 kgf (0.89 kgf to 0.09 kgf) vs −0.6 kgf (0.89 kgf to 0.2 kgf); 65.9 kgf (55.89 kgf to 76.47 kgf) vs 65.2 kgf (55.3 kgf to 75.8 kgf). Compared to model 1, model 2 provides minimally better results (Pseudo R^2^: 0.29 vs 0.28).

**Table 4.** Multivariable regression on generalized least squares, between handgrip test, social determinants of health and clinical parameters.

| **Model 1:** adjusted analysis for all predictors below |  |  |  |
| --- | --- | --- | --- |
| **Predictors (n=325)** | **Variance** | **CI 95%** | **P-value** |
| Age | -0.50 | 0.89 to 0.09 | 0.009 |
| Sex (male) | 65.86 | 55.89 to 76.5 | <0.001 |
| BMI | 0.40 | -0.09 to 1.0 | 0.13 |
| Number of medications (up to 3) | -2.57 | 8.51 to -3.7 | 0.41 |
| Number of medications (from 4 to 6) | -0.20 | 7.04 to -7.1 | 0.96 |
| Number of medications (7+) | -13.06 | 22.74 to 2.2 | 0.02 |
| **Pseudo R<sup>2</sup>: 0.28** |  |  |  |
| **Model 2:** adjusted analysis for significant variables in model 1 plus diabetes diagnosis |  |  |  |
| Age | -0.60 | 0.89 to 0.2 | 0.005 |
| Male sex | 65.2 | 55.3 to 75.8 | <0.001 |
| Number of medications (up to 3) | -1.88 | 7.8 to -4.6 | 0.56 |
| Number of medications (from 4 to 6) | -1.51 | -5.7 to 9.3 | 0.69 |
| Number of medications (7+) | -10.77 | 21.3 to -1.2 | 0.07 |
| Diabetes diagnosis (yes) | -2.76 | 10.1 to -5.0 | 0.47 |
| Diabetes diagnosis (no) | 2.94 | -14.3 to 23.7 | 0.75 |
| **Pseudo R<sup>2</sup>: 0.29** |  |  |  |

**Note:** 95%CI: Confidence Interval; BMI: Body Mass Index; R<sup>2</sup>: R Square. Estimation method: generalized least squares method. The reference category for sex: is female; number of medications: no medications and diabetes diagnosis: gestational diabetes.

## DISCUSSION

In the present exploratory study, we assessed whether predictors would contribute to predicting the 6MWT and handgrip test based on a conceptual framework construct. The main findings of our study are that social determinants of health and clinical parameters can have both direct and inverse associations with functional outcomes. Importantly, the addition of such well-identified explanatory variables (model 2) did not substantially modify the magnitude of effect, precision, and mean explanatory capacity of the outcome variance. Therefore, we infer that these variables did not present themselves as confounders nor modifiers of effect. While for 6MWT, age, sex (women), BMI, and QoL had a relevant mean clinical impact as predictors, only sex (men) impacted importantly handgrip strength, regardless of any statistical significance.

Several variables can impact the 6WMT performance, among them, height, age, sex, body weight are variables that can be highlighted because they are part of the reference equations for the distance covered in the test (ATS Committee on Proficiency Standards for Clinical Pulmonary Function Laboratories, 2002; Britto et al., 2013). Age, sex, and BMI are well-established predictors of 6WMT performance (ATS Committee on Proficiency Standards for Clinical Pulmonary Function Laboratories, 2002; Casanova et al., 2011; V. Z. Dourado, 2011; Salbach et al., 2015; Tveter et al., 2014) and the same was found in our study. Walking capacity is a proxy of QoL in a causal relationship (Lord & Menz, 2002; Serra et al., 2015). In his regard, we observed that for each additional score in the SF-6D questionnaire, around 21m was added to the walking capacity on average. Although clinically important, the cross-sectional nature and causal principles (such as Hill’s criteria for causality) (Hill, 1965) do limit a claim and put this independent relationship at risk of reverse causality. Lastly, the use of medications, diabetes diagnosis, depression, and family income did not explain any variability in the mean 6MWT values of the sample. While expected, this may have been an approximation of the QoL scores of our sample, which were roughly impaired at baseline, and consequently did not change the model.

Regarding the analyses using the handgrip strength as an outcome, sex and age were the preditors linked to the test performance, consistent with previous studies (Corrêa et al., 2020; Lu et al., 2020; Tveter et al., 2014). Diabetes did not predict changes in this outcome, although this disease leads to peripheral neuropathy and overall inflammation (Matsuda et al., 2004; Tsalamandris et al., 2019). To note, only 13% of the sample self-reported a diagnosis of diabetes and, additionally, we did not collect data regarding comorbidity related to diabetes (e.g., peripheral neuropathy). In both walking and handgrip strength tests, one possible contributing factor for diabetes not being a significant predictor may be the due the physical-active status of study participants. Some classes of medications may also impair strength production, such as antihypertensives and lipid-lowering drugs (Ashfield et al., 2010; Love et al., 2020; Onder et al., 2002), which were prevalent in our sample (F. M. Dourado et al., 2021). Nonetheless, BMI may be collinear to diabetes and reflect an inherent effect of disease, mirroring the effect of the disease in the model. After all, even for the variables which appeared to be correlated, the contribution to force magnitude was clinically marginal, except for sex, since being male explained to perform an average of 66.0 kgf more.

Our results should be interpreted in light of limitations. First, the mean values for our predicted outcomes (walking and handgrip) were similar to those of healthy and physically active ones, which may be a consequence of the engagement of our sample in a lifestyle program. Also, although using the conceptual framework model to choose explanatory variables with a solid rationale, some of them may present reverse causality in this study setting (cross-sectional study), regardless of our efforts to annulate this type of bias; also, we considered only one conceptual framework model for each multivariable analysis. The addition of more models and the comparability could allow us to better explain the phenomenon, which was diagnosed, at least statistically, by the presented R^2^ (around 30% of the mean variability of outcomes). For the handgrip strength variable, the original distribution was a Poisson-like distribution. As we decided to maintain betas to see contributions in real units, the variable was log-transformed. However, a sensitivity analysis using a robust variance Poisson regression with exponentialized betas (prevalence ratios - PRs) is a recommended future direction from our side.

On the other hand, we emphasize the exploratory analysis of correlates of these two outcomes, by its clinical relevance, particularly in this setting - in the older and majority in women. There is a pathophysiological rationale to assume a decline in the functional capacity during the aging process, here assumed as a combination of 6WMT performance and handgrip test performance in the older adults, due to impairments in various physiological systems of the body. To quantify the contribution of such parameters, as attenuators of contributors to the decline, is of importance, and we did this in a population-based study. Cautious should be taken into account for extrapolation in terms of sex, as a matter of importance. Then, our results should be considered for older people who are not healthy, but under treatment (pharmacological and non-pharmacological). Further exploration of the diagnosis of diabetes, number of medications, depression, and family income are also welcome.

## CONCLUSION

We conclude that in an older population engaged in a municipal program physical activity program, the walking capacity and handgrip strength were directly and inversely associated with social determinants of health and clinical parameters (sex, age, BMI, and QoL). The results may inform of programs or studies with comparable subjects cohorts and provide evidence for model adjustment in observational intervention studies targeting functional capacity in older subjects.

## Data Availability

All data produced are available online at (https://zenodo.org/record/4341443#.X9u61thKhPY; doi: 10.5281/zenodo.4341443).

https://osf.io/q4r69/

## Acknowledgments

We would like to thank Rogério Boff Borges, a statistician at the Biostatistics Unit of the Hospital de Clínicas of Porto Alegre, for his help and guidance in carrying out the statistical analysis for this article.

